# COVID-19 in children with rheumatic diseases (RD) in the spanish national cohort EPICO-AEP

**DOI:** 10.1101/2020.10.17.20214296

**Authors:** Cristina Calvo, Agustín Remesal, Sara Murias, Fátima Ara-Montojo, Enrique Otheo, Francisco J Sanz-Santaeufemia, Álvaro Villarroya, Clara Udaondo, Cinta Moraleda, Alfredo Tagarro, on behalf of EPICO-AEP Working Group

## Abstract

**Objectives:** SARS-CoV-2 infection in adults with rheumatic diseases (RD) is a cause for concern. Data in the pediatric population are practically absent. We aimed to describe the prevalence of patients with RD and their complications among children admitted with COVID-19 in the Spanish national cohort EPICO-AEP; a multicenter prospective national study.

**Methods:** Children <18 years old with RD and COVID-19 enrolled in EPICO-AEP were included in this study.

**Results:** By June 30th 2020, 350 children were admitted in secondary and tertiary hospitals of Spain with SARS-CoV-2 infection. A total of 8 patients presented RD (2.2% of those hospitalized). All were female. The median age was 12.1 years (IQR 8.3-14.5). The diagnosis related with COVID-19 were febrile syndrome and/or upper respiratory infection (4 cases) and pneumonia (4 cases). One of the 8 (12.5%) patients with a severe juvenile dermatomyositis (JDM) with interstitial lung disease died. Juvenile idiopathic arthritis (JIA) was the most frequent diagnosis in 3/8 (37.5%) patients. In 5/8 (62.5%) cases, the RD was not fully controlled, and all patients except one received corticosteroid treatment.

**Conclusions:** Children with RD have accounted for 2.2% of hospitalized patients with COVID-19 in our series. The evolution has been moderately favorable, with one deceased. In case of active disease and use of corticosteroids patients should be managed with caution.

**What is already known about this subject?:**

- Studies in adults with rheumatic disesases (RD) show that immune-mediated inflammatory disease and use of biologics are not associated with worse COVID-19.
- Poorly controlled active RD or some treatments such as corticosteroids, may have an increased risk of infection and serious disease in adults. No data in children.

**What does this study add?:**

- Pediatric data from the national EPICO-AEP registry in Spain are presented, where children with RD and COVID-19 were 2.2% of those hospitalized.
- Active disease and the use of corticosteroids could be considered risk factors in the pediatric population as well as in adults.

**How might this impact on clinical practice or future developments?:**

- The rheumatologist pediatrician must be very careful in the management of children with COVID-19 infection, especially if they receive corticosteroid treatment or have active RD.

## INTRODUCTION

SARS-CoV-2 infection in children is relatively mild, although the pediatric population suffers at least 10% of identified cases[1] with a small proportion of them requiring hospitalization. From 25% to 60% of admitted children with COVID-19 have comorbidities, including rheumatic diseases (RD)[2,3]. Pediatric patients with RD often receive immunosuppressive medication and in theory, they are a group of risk.

Studies in adults with RD show that immune-mediated inflammatory disease and use of biologics are not associated with worse COVID-19 [4-7]. However, if patients have poorly controlled active RD or receive some treatments such as corticosteroids, they may have an increased risk of infection and serious disease. Based on the results of the published series, some scientific associations have relased management recommendations for these patients[8]. The American College of Rheumatology (ACR) has made recommendations for SARS-CoV-2 infections in children with RD, but they acknowledge the lack of pediatric data[9].

We aimed to describe the prevalence of patients with rheumatic diseases and their complications among children admitted with COVID-19 in the Spanish national cohort EPICO-AEP.

## PATIENTS AND METHODS

The Epidemiological Study of COVID-19 in Children of the Spanish Society of Pediatrics (EPICO-AEP) is a multicenter prospective national study aiming to describe the pediatric COVID-19 in Spain. Children younger than 18 years with infection due to SARS-CoV-2 and attended at 49 hospitals were included in this registry. Inclusion criteria included positivity in real-time polymerase chain reaction (RT-PCR) in nasopharyngeal sample, IgM or IgG in lateral-flow rapid test, ELISA or chemiluminescence serology, or severe disease suggestive of multi-Inflammatory syndrome related to SARS-CoV-2 (MIS-C) and recent household contact with a confirmed patient with COVID-19. From March 1st to June 30th, 2020, children with RD and COVID-19 enrolled in EPICO-AEP were included in this study.

This study was approved by the ethics committee of Hospital 12 de Octubre (code 20/101). Informed consent was obtained by parents and mature minors. Data were analyzed using the Stata version 15, College Station, TX.

## RESULTS

By June 30th 2020, 498 children attended in secondary and tertiary hospitals of Spain were diagnosed with SARS-CoV-2 infection and included in the EPICO-AEP cohort (manuscript in preparation). The median age was 61 months (Interquartile range (IQR) 6-140) and 54.6% were males. Of those, 350 (72.9%) were hospitalized and 262 (52.6%) had a symptomatic COVID-19 disease, 48/350 (13.7%) patients requiring intensive care unit admission, and 4/350 children died (1.1% of hospitalized).

A total of 8 patients presented RD (2.2% of those hospitalized). All were female and the median age was 12.1 years (IQR 8.3-14.5). The clinical data are shown in Table 1. The admission diagnosis related with COVID-19 were febrile syndrome and/or upper respiratory infection in 4 cases and pneumonia in 4 cases (one of them with respiratory failure). Unfortunatly, 1 of the 8 patients with RD (12.5%) died. She was 1 of the 4 only patients (25%) deceased within the whole cohort. She had a severe juvenile dermatomyositis (JDM) with interstitial lung disease.

**Table 1.** Clinical characteristics of children with rheumatic diseases and SARS-CoV-2 infection.

| **Case #** | **1** | **2** | **3** | **4** | **5** | **6** | **7** | **8** |
| --- | --- | --- | --- | --- | --- | --- | --- | --- |
| **Age (years)** | > 10 | > 10 | >10 | < 10 | >10 | <10 | >10 | >10 |
| **Gender** | F | F | F | F | F | F | F | F |
| **COVID-19 close contact** | Yes | No | No | No | Yes | No | Yes | Yes |
| **Primary disease** | ‡JDM-MDA5 with interstitial lung disease | C-ANCA vasculitis. Hemodialysis | SJIA | SJIA | JIA Oligoarticular | PFAPA | Polyarteritis nodosa | Antiphospholipid syndrome. Suspicion of SLE |
| **Immunosuppressive treatment/targeted therapy** | `Metilprednisolone pulses, tacrolimus, MMF, cyclophosphamide immunoglobulins. | Prednisone (5 mg/day), azathioprine | Metilprednisolone pulses followed by prednisone 1 mg/kg/day | Prednisone 1 mg/kg/day, canakinumab | MTX, infliximab | Prednisone intermittent (2 mg/kg/ single dose when needed; not recently) | Prednisone 2.5 mg/48 hours, MTX, colchicine, tocilizumab | Prednisone 0,5 mg/kg/day |
| **Time since RD diagnosis** | 4 months | 1 year | Onset | 2 months | 10 years | 2 years | 6 years | Onset |
| **Fever** | Yes | Yes | Yes | Yes | Yes | Yes | No | Yes |
| **Cough** | Yes | Yes | Yes | Yes | No | Yes | Yes | No |
| **Sore throat** | Yes | Yes | Yes | No | Yes | No | No | No |
| **Dyspnea** | Yes | No | No | No | No | Yes | No | No |
| **Lymphocytes/mm<sup>3</sup> (minimum value)** | 0 | 380 | 1070 | 1080 | 2950 | 2300 | 2900 | 1200 |
| **D-dimer ng/mL (maximum value)** | 10200 | 1168 | 829 | 5953 | 142 | - | 174 | 1194 |
| **IL-6 pg/mL (maximum value)** | 1000 | - | - | - | - | - | - | - |
| **C- Reactive Protein (mg/L)** | 41,4 | 9 | 103,8 | 341 | 3 | 12,1 | 0,3 | 33 |
| **Ferritin (mg/dl)** | 60456 | 1270 | 419 | 5200 | - | - | 84 | 517 |
| **Chest X-ray** | Bilateral infiltrate.<br>Severe interstitial<br>lung disease<br>Pneumothorax | Normal<br>initially but<br>focal infiltrate<br>on evolution | Subtle ground-glass<br>infiltrate in the left<br>lung base,<br>asymmetrical | Lobar<br>consolidation<br>pleural effusion | Normal | Bilateral<br>infiltrate | Normal | Normal |
| **Required**<br>**hospitalization** | Yes | Yes | Yes | Yes | Yes | Yes | Yes | Yes |
| **Oxygen therapy** | Yes | Yes | No | Yes | No | No | No | No |
| **Treatment** | Antibiotics<br>HCQ<br>Remdesivir**<br>Tocilizumab | Reduction in<br>IS<br>(azathioprine<br>withdrawal)<br>LPV/Rtv<br>HCQ* | HCQ | LPV/Rtv<br>Antibiotics | - | Antibiotics | HCQ | HCQ<br>Enoxaparin |
| **Duration of**<br>**hospitalization (days)** | 33 | 7 | 9 | 12 | 3 | 6 | 2 | 30 |
| **Complications** | Sepsis/Shock<br>Respiratory & Renal<br>Failure<br>Mechanical<br>ventilation<br>Decease | None | None | Femoral vein<br>thrombosis<br>catheter<br>related.<br>Suspicion of<br>bacterial<br>infection | None | None | None | Femoral vein<br>thrombosis.<br>Adrenal<br>hemorrhage |
| **Seroconversion** | - | Yes | - | No | No | - | Yes | - |

F: female, JDM: Juvenile dermatomyositis; HCQ: hydroxychloroquine, IS: immunosuppressors, JIA: Juvenile Idiopathic Arthritis; SJIA: Systemic Juvenile Idiopathic Arthritis; LPV/Rtv: lopinavir/ritonavir, O2: oxygen; MMF: mycophenolate; MTX: methotrexate; RD: rheumatic disease; SLE: Systemic lupus erythematosus
\*HCQ According to our local guidelines at admission, the oral dose was as follows: HCQ 6.5 mg/kg/day (dosing q12 hours) in >6 year-olds, and HCQ 10 mg/kg/day (dosing q12 hours) in children > 6 years (maximum daily dose 400mg) for 5 days.
\*\* Remdesivir was given for a total of 5 days at an intravenous dose of 5 mg/kg on day 1, followed by a maintenance dose of 2.5 mg/kg.

Juvenile idiopathic arthritis (JIA) was the most frequent diagnosis, and was present in 3/8 (37.5%) patients. In 5/8 (62.5) cases, the RD was not fully controlled and had been diagnosed <1 year before. In 2 patients, the diagnosis of COVID-19 was coincident in time with the onset of RD. In the most severe case, who had juvenile dermatomyositis (JDM) with interstitial lung involvement, the disease was always active and she did not evolve favorably since the diagnosis. Although 7 of the cases had a good outcome, complications included 2 deep vein thromboses and 1 pulmonary bacterial superinfection. All patients except one received corticosteroid treatment for their underlying condition (although 1 patient with PFAPA syndrome only received steroids intermittently). Other immunosuppressants are detailed in Table 1. Immunosuppressive treatment was not fully suspended in any case (azathioprine was withdrawn in one case). Three patients required oxygen, and most (6/8; 75%) were treated with hydroxychloroquine or lopinavir/ritonavir as recommended by local guidelines at the time of admission. Only the most serious case received remdesivir.

Special mention deserves the patient with JDM, that has been previously published[10]. Anti-melanoma differentiation-associated gene 5 juvenile dermatomyositis (anti-MDA5 JDM) is associated with a high risk of developing rapidly progressive interstitial lung disease. She was complicated with a secondary hemophagocytic lymphohistiocytosis and during her clinical course evolution a RT-PCR for SARS-CoV-2 performed in bronchial aspirate sample was positive. Despite the aggressive treatment of interstitial lung disease due to anti-MDA5 JDM, there was no improvement of respiratory failure in the following days and the patient developed refractory septic shock and died.

## DISCUSSION

We report the prevalence of children with COVID-19 and RD in the Spanish national EPICO-AEP cohort of hospitalized children. As far as we know, it is the first national cohort that collects children with RD, representing 2.2% of admitted children with COVID-19 in Spain. Although the general outcome was favorable, 1 patient (12.5%) died and 2 had thrombotic complications. Most of the patients received prednisone, which has been considered a drug that confers an increased risk of severe COVID-19 in adults with RD[4,5].

The series published in adults agree that patients with RD have a similar risk to the general population of contracting a SARS-CoV-2 infection and those having other comorbidities like hypertension, diabetes or cardiac diseases are at higher risk of developing a more severe course of COVID-19[4-7]. However, certain conditions such as the presence of active disease, and some immunosuppressants confer an increased risk of hospitalization. Thus, in the German National register in 104 adults with RD and COVID-19, hospitalized patients were more often treated with glucocorticoids while biological disease-modifying antirheumatic drugs (DMARDs) were used less often[4]. The Global Rheumatology Alliance physician-reported registry[5], with a total of 600 cases of COVID-19 in adults with RD from 40 countries, found that nearly half of the cases were hospitalized (277, 46%) and 55 (9%) died. In a multivariable-adjusted model, prednisone dose ≥10 mg/day was associated with higher odds of hospitalization. The use of conventional DMARD alone or in combination with biologics/Janus Kinase inhibitors was not associated with hospitalization. Neither does the use of non-steroidal anti-inflammatory drugs (NSAID) was associated with hospitalization. Tumour necrosis factor inhibitor (anti-TNF) use was associated with reduced odds of hospitalization, while no association with hydroxychloroquine use was observed. Other adult’s Spanish series[6] with 122 patients with RD found that treatment with methotrexate (MTX) and rituximab was a risk factor for hospital admission, but not for mortality, while other DMARDs did not show differences. However, glucocorticoids seem to increase the risk of mortality.

Chronic steroids have a strong immunosuppressive effect, as long as hypothalamic-hypophysis axis suppression. Our results, along with literature in adults, suggest that steroids might have more risk for COVID-19 admission than biologics. Pediatricians should be cautious when evaluating children on chronic steroids and COVID-19.

For children with RD and COVID-19, the ACR[8] recommends maintaining treatment with NSAID, conventional DMARDs and biological DMARDs. Regarding glucocorticoids, the task force recommended that glucocorticoids should be continued or initiated when clinically indicated, using the lowest effective dose to control underlying RD without defining a specific dose (e.g., less than or equal to 10mg prednisone used in adults) for the pediatric population. In our patients, the treatment was not modified, except for azathioprine withdrawal in one case. Most patients had active disease, and the risk of withdrawal was considered higher than the risk of maintaining the drugs. Although more data in children with RD are necessary to release strong recommendations, our series seems to support that corticosteroid treatment may be a risk factor for hospitalization in this group of children.

Data in the pediatric population are practically absent. In 2 series from Italy and UK, COVID-19 in children with rheumatic diseases were not reported[3,11]. In our series, it is worth mention that 100% were females and age was higher than the overall cohort, possibly reflecting the pediatric population of patients with RD. Virtually all patients were receiving treatment with prednisone or MTX that have been found to risk hospitalization or serious illness in adults[6]. Importantly a significant percentage was in the early stages of the disease, without good control of it. Two patients had COVID-19 at the time of onset of RD, which raises the hypothesis if COVID-19 may have a role triggering the disease. This hypothesis have also been formulated for other autoimmune disease, as diabetes[12]. Overall outcome was good, except in the case of JDM, and interstitial lung disease has been recognized as risk factor of severe COVID-19 in adults[13]. Thrombotic complications are common in adults with COVID-19 but are rare in children. In our series two girls presented them; one of them related to central venous catheter and the other in the context of antiphospholipid syndrome and suspicion of systemic lupus erythematosus (SLE), so the role that SARS-CoV-2 infection can play in this complication is uncertain.

In summary, in the Spanish national EPICO-AEP cohort, children with RD have accounted for 2.2% of hospitalized patients with COVID-19. The evolution has been moderately favorable, with one deceased. Active disease and the use of corticosteroids could be considered risk factors in the pediatric population as well as in adults. More prospective studies are needed to characterize risk factors in this population.

## Data Availability

Data is available on request

